# Adequacy of Nasal Self-Swabbing for SARS-CoV-2 Testing in Children

**DOI:** 10.1101/2022.03.07.22270699

**Authors:** Jesse J. Waggoner, Miriam B. Vos, Erika A. Tyburski, Phuong-Vi Nguyen, Jessica M. Ingersoll, Candace Miller, Julie Sullivan, Mark Griffiths, Cheryl Stone, Macarthur Benoit, Laura Benedit, Brooke Seitter, Robert Jerris, Joshua M. Levy, Colleen S. Kraft, Sarah Farmer, Amanda Foster, Anna Wood, Adrianna L. Westbrook, Claudia R. Morris, Usha N. Sathian, William Heetderks, Li Li, Kristian Roth, Mary Barcus, Timothy Stenzel, Greg S. Martin, Wilbur A. Lam

**Affiliations:** Emory University School of Medicine, Atlanta, GA, USA; Children’s Healthcare of Atlanta, Atlanta, GA, USA; Georgia Institute of Technology, Atlanta, GA, USA; Pediatric Biostatistics Core, Department of Pediatrics, Emory University; Consultant, National Institute of Biomedical Imaging and Bioengineering, NIH, Bethesda, MD; Division of Microbiology, OHT7 Office of Product Evaluation and Quality, Center for Devices and Radiological Health, U.S. Food & Drug Administration; OHT7 Office of Product Evaluation and Quality, Center for Devices and Radiological Health, U.S. Food & Drug Administration

## Abstract

**Background:** The goal of this study was to characterize the ability of school-aged children to self-collect adequate anterior nares (AN) swabs for severe acute respiratory syndrome coronavirus 2 (SARS-CoV-2) testing.

**Methods:** From July to August 2021, 287 children, age 4-14 years-old, were prospectively enrolled in the Atlanta area. Symptomatic (n=197) and asymptomatic (n=90) children watched a short instructional video before providing a self-collected AN specimen. Health care workers (HCWs) then collected a second specimen, and useability was assessed by the child and HCW. Swabs were tested side-by-side for SARS-CoV-2. RNase P RNA detection was investigated as a measure of specimen adequacy.

**Results:** Among symptomatic children, 87/196 (44.4%) tested positive for SARS-CoV-2 by both self- and HCW-swab. Two children each were positive by self- or HCW-swab; one child had an invalid HCW-swab. Compared to HCW-swabs, self-collected swabs had 97.8% and 98.1% positive and negative percent agreements, respectively, and SARS-CoV-2 Ct values did not differ significantly between groups. Participants ≤8 years-old were less likely than those >8 to be rated as correctly completing self-collection, but SARS-CoV-2 detection did not differ. Based on RNase P RNA detection, 270/287 children (94.1%) provided adequate self-swabs versus 277/287 (96.5%) HCW-swabs (p=0.24) with no difference when stratified by age.

**Conclusions:** Children, aged 4-14 years-old, can provide adequate AN specimens for SARS-CoV-2 detection when presented with age-appropriate instructional material, consisting of a video and a handout, at a single timepoint. These data support the use of self-collected AN swabs among school-age children for SARS-CoV-2 testing.

## Introduction

As the severe acute respiratory syndrome coronavirus 2 (SARS-CoV-2) pandemic has evolved, testing has become widely available for children and adults who may have coronavirus disease (COVID-19), the disease caused by SARS-CoV-2. However, testing capacity remains insufficient for repeat testing of asymptomatic school children and group settings for children, such as camps and schools, that are less likely to have trained health care workers (HCWs) available for sample collection. Long-standing medical practice has been to have a HCW collect samples for respiratory tract infection testing, which typically involves swabbing in the nose or mouth. Little data exists to suggest that HCW-collection is necessary, and it remains a barrier to expanding testing in locations with large numbers of children and limited access to healthcare personnel. Comparison of parental and nurse nasal swab collection for respiratory tract infection in children has demonstrated that nasal swab samples collected by parents are comparable to nurse-collected samples,^1^ however even this has not become routine for younger children.

Many SARS-CoV-2 tests have been authorized by the United States Food and Drug Administration (FDA) under Emergency Use Authorizations (EUAs) for self-swabbing by adults and children ≥14 years-old, and standardized instructions for self-swabbing of anterior nares (AN) are available. Many tests are also authorized by the FDA under EUA for parental swab sample collection for children ≥2 years-old. AN swabs are preferred for self-collection because they are technically less complex. However, the age at which nasal self-swabbing would generally be successful and how much it may affect sensitivity of COVID-19 testing is not known, and tests authorized by the FDA under EUA are not authorized for self-swabbing by children <14 years-old. Thus, there is a need to better understand at what age children can self-swab to provide a specimen for testing. This study was a collaboration between Emory University, Children’s Healthcare of Atlanta, the Rapid Acceleration of Diagnostics (RADx) initiative of the National Institutes of Health National Institute of Biomedical Imaging and Bioengineering (NIH, NIBIB), and the FDA to generate data to support evidence-based recommendations to schools and other group settings with children regarding utilization of self-swabbing. Because the definition of an adequate sample was undefined from our review of the literature, we conducted a multi-part study to answer the following study questions: 1) Are children, after hearing and seeing simple instructions, able to self-collect AN swabs for SARS-CoV-2 testing? 2) In children with suspected COVID-19, is self-swabbing as effective for detecting SARS-CoV-2 as HCW-swabbing? And 3) Can RNase P RNA serve as an indicator of AN swab specimen adequacy during periods of low respiratory virus transmission?

## Methods

### Ethics statement

All studies were approved by the Emory University IRB and all participants provided consent (and assent, where indicated by age) prior to participating.

### Symptomatic children

A prospective cohort of symptomatic SARS-CoV-2 positive and negative children was recruited into a cross sectional study to compare self-swabbing to HCW-swabbing for SARS-CoV-2 detection. Symptomatic children were recruited based on a daily report generated of all children testing positive by standard of care nasopharyngeal (NP) swab within the Children’s Healthcare of Atlanta System in the previous 24 hours. Target enrollment was 10 COVID-positive and 10 COVID-negative children in each age group. Parents were called and asked to return to a Drive-Thru testing site set up at the Center for Advanced Pediatrics. Children were excluded if self-swabbing was not feasible due to a medical condition; developmental delay precluded the child from understanding the instructions in the opinion of the parent (or guardian); or the child had a history of nosebleeds in the past 2 weeks. If a SARS-CoV-2-positive child returned for the self-swabbing study ≥48 hours after their most recent positive test, a repeat, standard-of-care NP COVID test was obtained after AN swab collection for the current study.

Once consented, a video of children teaching and demonstrating how to self-swab was shown, and children were provided with an instructional handout with images on one side and images plus written instructions on the other (Supplemental Material). The child then performed the self-swab followed by the HCW. The instructions in the video and handout were used as the process for swabbing. Four rotations of a Nylon Flocked Swab (regular size with 30mm breakpoint; Copan Diagnostics, Murrieta, CA) were performed in each naris. Then the swab was given to the HCW, who placed it in a sterile cryovial pre-filled with 1mL saline. The same process was repeated by the HCW. The 3 HCWs who conducted all swabbing for this project were highly skilled and experienced pediatric nurses. Following collection, samples were immediately placed on ice and transported to the laboratory by courier at the end of each day. Samples were stored at 4°C for up to 72 hours prior to nucleic acid extraction and testing. If the expected duration of storage exceeded 72 hours, samples were stored at -80°C.

### RNase P RNA detection – Pilot study

The overall study was planned and initiated during the nadir in SARS-CoV-2 transmission that occurred during the spring and early summer of 2021.^2^ As such, RNase P RNA detection was evaluated as an investigational measure of specimen adequacy. A retrospective pilot study was conducted to determine if RNase P RNA could be detected in HCW-collected AN samples. A set of 24 archived AN samples from asymptomatic children age 5 to 17 years were sourced from the Children’s Clinical and Translational Discovery Core, then re-extracted and tested using the investigational study protocol for RNase P RNA detection. Selected samples included 12 without detectable SARS-CoV-2 RNA and 12 with detectable SARS-CoV-2 RNA grouped by cycle threshold (Ct): 4 samples each having Cts of <24, 24-32, and >32. All had been stored at -80°C in normal saline until extraction.

### Asymptomatic children

Following completion of the pilot study evaluating RNase P RNA detection, asymptomatic children were prospectively enrolled, with a target enrollment of 10 children per year of age from 5 to 12 years. Recruitment was conducted through a metro Atlanta neighborhood social media page. Parents were invited to sign their child/children up for 15-minute time slots over the course of 2 days. An outdoor testing tent was set up near the neighborhood clubhouse. Children came with their parents and consent and assent were obtained. A small number of children were recruited by word of mouth to target specific ages that were needed to achieve the target enrollment following these 2 days. In addition to the exclusion criteria for symptomatic children, asymptomatic children were also excluded if COVID-19 had been suspected/diagnosed in the past 30 days or if the child had current respiratory symptoms or other COVID-19 symptoms in the past 2 weeks. Children observed the same video and were provided with the same instructional handout prior to self-swabbing, as described above. The research team of nurses and research coordinators followed the same process as described for the symptomatic cohort.

### Usability evaluation

A set of usability questions was developed with the goal of determining the feasibility and overall experience of self-swabbing by children (Supplemental Material). The aim of the questions was to determine if all tasks associated with self-swabbing were completed correctly, whether participants required any assistance, how participants felt about self-swabbing, and whether participants understood the instructions. A portion of the questions were HCW-facing; that is, the HCW conducting the study observed participant actions and responded to questions accordingly. The remaining questions were participant-facing. The usability questionnaire was completed immediately after sample collection by the child and HCW. The child received assistance from the HCW, as needed, to complete the usability questionnaire.

Given that the HCWs administering the questions were also focused on conducting the study, the data entry required was intended to be minimally disruptive. The participant-facing questions came with a separate set of considerations, as the participants were children. The vocabulary used in the questions was selected to be accessible to the age groups included in the study. Questions were designed to be non-leading and open-ended where possible, and used prompts such as “can you tell me about….,” as these methods have been show to elicit more complete and accurate information from children.^3,4^ Questions were designed to be answered with a *yes/no* selection, selection from a list of options, or, for participant-facing questions, one to two words summing up responses to open-ended questions.

### SARS-CoV-2 molecular testing

All samples from symptomatic participants and the single asymptomatic participant with SARS-CoV-2 were extracted and tested with the CDC 2019-Novel Coronavirus (2019-nCoV) Real-Time RT-PCR Diagnostic Panel (hereinafter referred to as the CDC EUA rRT-PCR) according to the instructions for use.^5^ Briefly, 100µL of sample was extracted on a Roche MagNA Pure 96 using the DNA and Viral NA Small Volume Kit (Roche, Basel, Switzerland). Nucleic acids were eluted in 100µL and immediately tested for the N1, N2 and RNase P in separate 20uL reactions of the TaqPath 1-Step RT-qPCR Master Mix (Thermo Fisher, Waltham, MA) on an ABI Fast DX Real-Time PCR system (Thermo Fisher). Results were interpreted according to the instructions for use.^5^ All samples with inconclusive results were re-tested by rRT-PCR, and samples with invalid results were re-extracted and re-tested.

### Investigational RNase P RNA detection

All samples were tested for SARS-CoV-2 and RNase P RNA with an investigational study protocol. Total nucleic acids were extracted from 200µL of AN sample and eluted into 50µL on an EMAG instrument (bioMérieux, Durham, NC). Following extraction, 12µL of eluate was DNase treated in 25µL reactions of the Heat&Run gDNA Removal Kit (ArticZymes, Tromsø, Norway) according to manufacturer recommendations. Resulting RNA was immediately tested by real-time RT-PCR (rRT-PCR) for the nucleocapsid 2 (N2) target and RNase P. The assay was run as duplex test but otherwise performed as described.^6^ A specimen was considered to have detectable SARS-CoV-2 or RNase P RNA if it yielded a Ct value ≤ 40 for that target in the duplex assay. Samples from 21 children with detectable SARS-CoV-2 RNA were also tested in a multiplex rRT-PCR for specific *spike* mutations associated with variants of concern. This assay was performed, as described,^7,8^ using DNase-treated RNA.

### Statistical analysis

For analysis of test outcomes, concordant pairs were defined as specimen pairs that generated the same qualitative result with the CDC EUA rRT-PCR. Negative concordant pairs had undetectable SARS-CoV-2 RNA but detectable signal for RNase P. Positive concordant pairs had detectable SARS-CoV-2 RNA. Useability data were evaluated by age and when binned as ≤8 and >8 years-old, based on initial review of the results. Descriptive statistics for the study were reported as medians and interquartile ranges for continuous variables and counts with percentages for categorical variables. Shapiro-Wilk tests were used to check normality of continuous data. Two-group comparisons were conducted using students t-tests for normally distributed continuous data. Wilcoxon rank-sum tests were used for non-normal continuous data. Categorical data was compared using chi-squared tests or Fisher’s exact tests for expected cell counts <5. Statistical significance was assessed at the 0.05 level. All statistical analyses were conducted using SAS 9.4 (Cary, NC).

## Results

### Symptomatic children

197 symptomatic children, aged 4-14 years-old, were enrolled in July-August 2021 (Figure 1). A single HCW-collected swab was invalid (no detectable SARS-CoV-2 or RNase P RNA), and this participant was removed from further analysis (self-swab, negative). 87 children (44.4%) tested positive for SARS-CoV-2 by both self- and HCW-swabs (positive concordant samples), and two children each tested positive by self- or HCW-swab, but negative by the alternate swab. Positive and negative percent agreements were 97.8% and 98.1%, respectively (kappa = 0.96, 95% CI: 0.92 to 0.99; Figure 2A). Children with concordant positive SARS-CoV-2 rRT-PCR results presented, on average, one day later than children who tested negative [days post-symptom onset, median 3 (IQR 1-4) vs 2 (1-3), respectively; p=0.002]. These two groups were otherwise similar (Table 1). N2 Ct values obtained with the CDC EUA rRT-PCR did not differ between self- and HCW-collected swabs among the 87 children with SARS-CoV-2 detected in both samples (Figure 2B).

**Figure 1.**
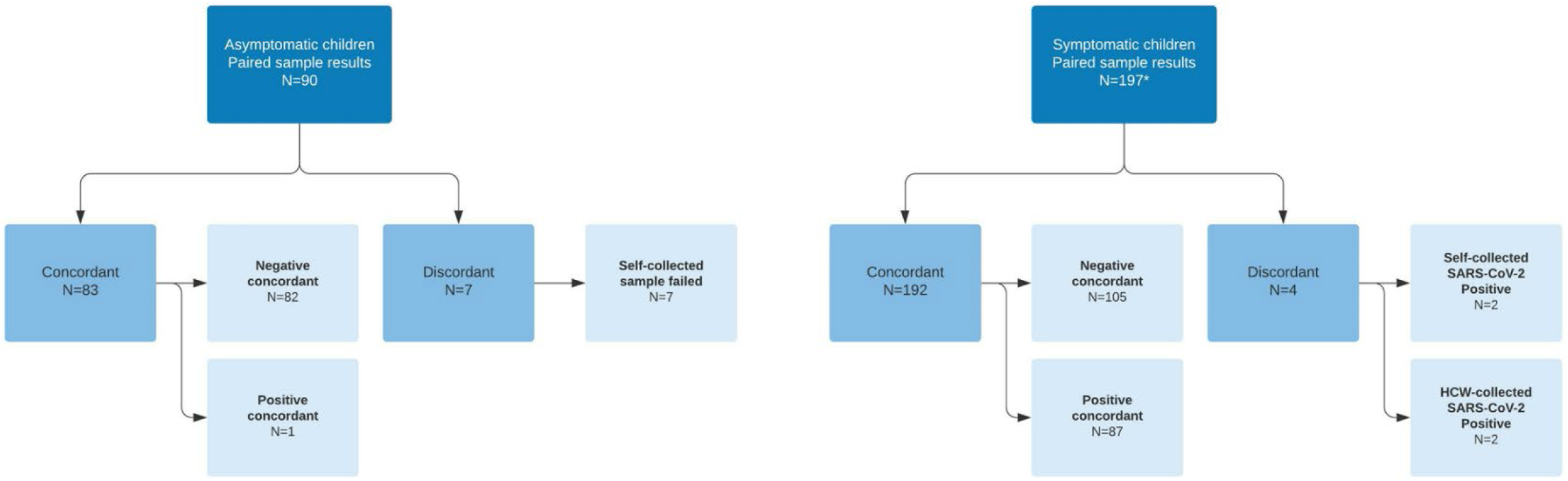
Study flowchart for samples from prospectively enrolled asymptomatic and symptomatic children. Self- and HCW-collected samples that yielded the same qualitative rRT-PCR result were considered concordant. * One child tested self-swab negative and HCW-swab invalid (no detectable RNase P).

**Figure 2.**
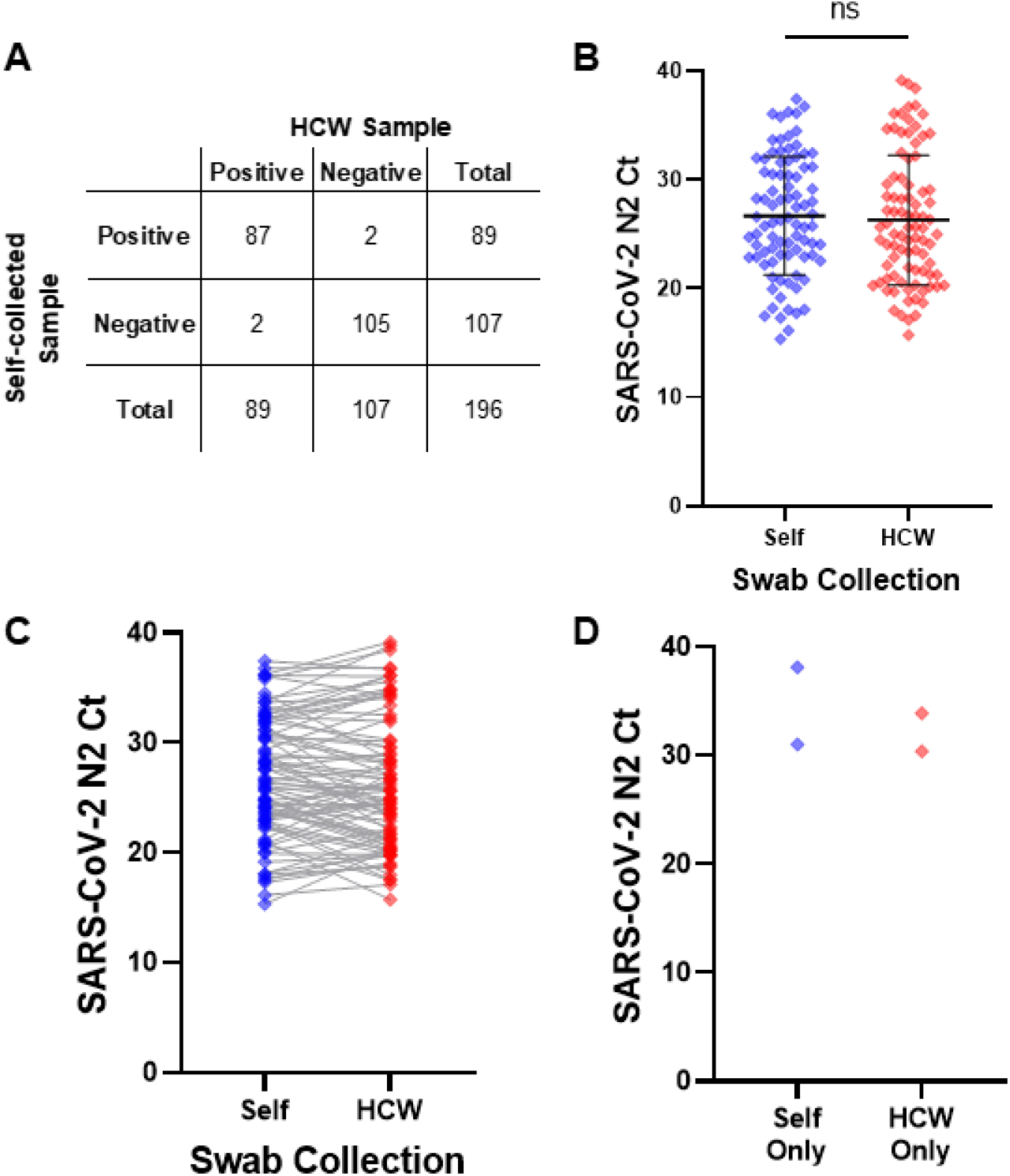
SARS-CoV-2 RNA detection does not differ between self- and HCW-collected nasal swabs in symptomatic children. **A**) 2×2 comparison of qualitative SARS-CoV-2 detection, **B**) overall distribution of SARS-CoV-2 N2 target Ct values, and **C**) paired SARS-CoV-2 N2 Ct values for self- and HCW-collected swabs. **D**) Ct values for the positive sample from 4 discrepant sample pairs where SARS-CoV-2 RNA was only detectable in the self-collected (n=2) or HCW-collected (n=2) specimen. All displayed results were obtained with the CDC EUA rRT-PCR.

**Table 1.**
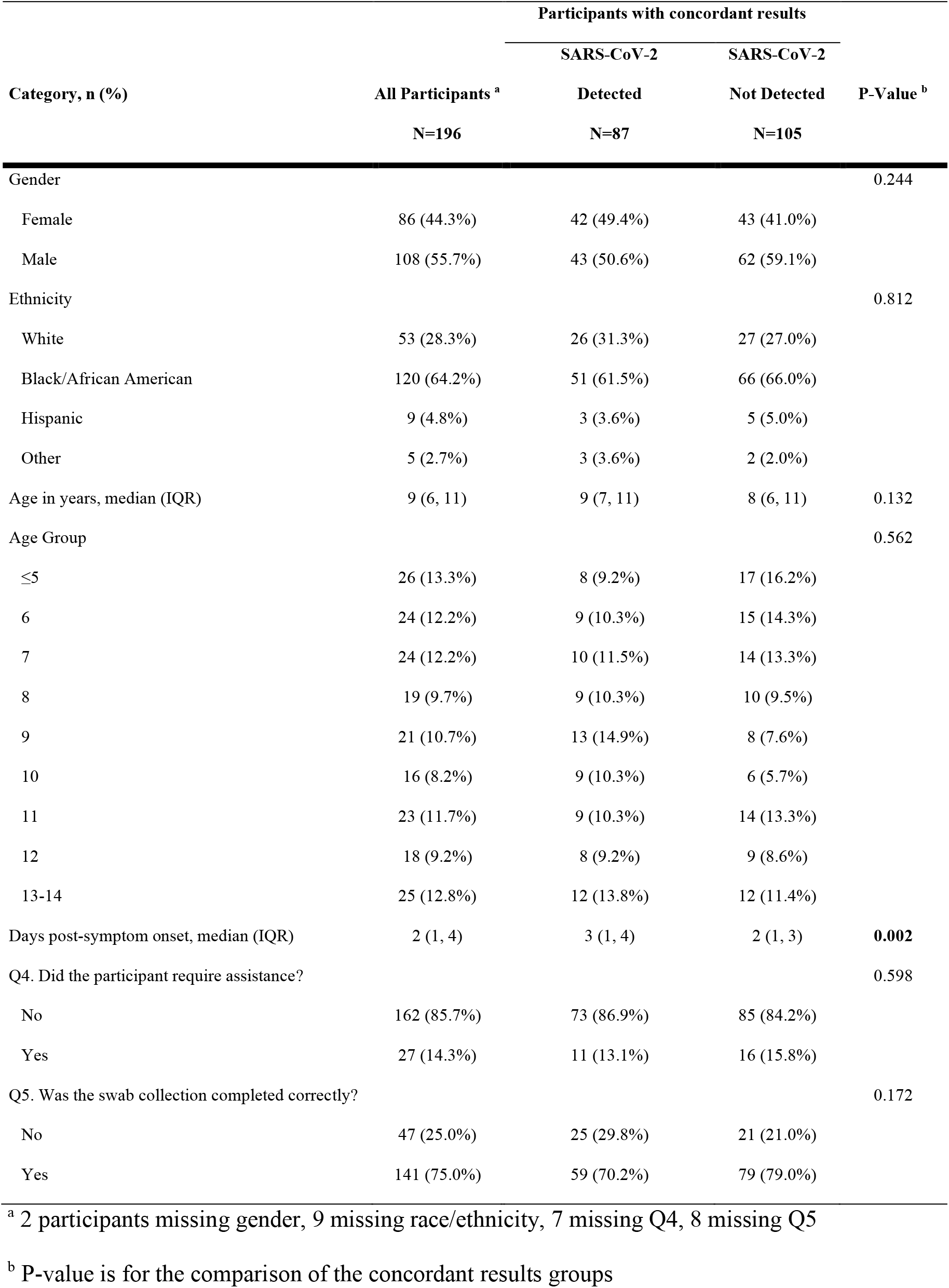
Characteristics of symptomatic children with concordant SARS-CoV-2 positive vs. SARS-CoV-2 negative samples.

For 28 participants (32.9%), N2 Ct values differed by > 3.3 cycles between self- and HCW-collected swabs, consistent with ∼10-fold difference in viral RNA concentration. 12 children (14.1%) had lower Ct values (greater RNA) in self-collected swabs and 16 (18.8%) had lower Ct values in HCW-collected swabs (Figure 3C). Children with lower Ct values in the self-collected swab were more likely to be black/African American and present earlier in the course of illness. No other differences were noted between these groups (Table S1). Finally, 4 participants had discordant qualitative SARS-CoV-2 results; all had N2 Ct values > 30.0 from the positive sample (Figure 2D, Table 2).

**Table 2.** Characteristics of participants with discordant SARS-CoV-2 results between self- and HCW-collected AN swabs tested in the CDC EUA rRT-PCR.

| Positive swab | SARS-CoV-2<br>N2 Ct | Age | Gender | Race | Ethnicity | Did the participant<br>require assistance? | Was the self-swab<br>collection completed<br>correctly? |
| --- | --- | --- | --- | --- | --- | --- | --- |
| *Self-collected swab* |  |  |  |  |  |  |  |
| Patient 8232 | 31.02 | 12 | Male | Black/African American | Non-Hispanic | No | Yes |
| Patient 8298 | 38.10 | 14 | Male | Black/African American | Non-Hispanic | No | Yes |
| *HCW-collected swab* |  |  |  |  |  |  |  |
| Patient 8113 | 30.38 | 10 | Male | Black/African American | Non-Hispanic | No | Yes |
| Patient 8244 | 33.87 | 4 | Female | Patient's race missing | Hispanic | No | No |

### Investigational RNase P RNA detection

In the retrospective pilot study, RNase P was detected in AN swabs from all 24 asymptomatic children. RNase P Cts were significantly higher in DNase treated versus untreated eluates (mean Ct 31.6 ± 2.7 vs. 23.8 ± 2.6, respectively; p <0.001, Figure S1A). RNase P RNA Cts did not differ significantly between samples with and without detectable SARS-CoV-2 RNA or based on the SARS-CoV-2 Ct in positive samples (Figure S1B and C). Based on the pilot data, 90 asymptomatic children, aged 5-12 years-old, were enrolled in July 2021 (Figure 1), and all self- and HCW-collected swabs from symptomatic and asymptomatic children were tested with the investigational study protocol.

For this phase of the study, an adequate specimen was defined as a specimen with detectable RNase P or SARS-CoV-2 RNA by rRT-PCR. Overall, 270/287 children (94.1%) provided adequate self-swabs versus 277/287 (96.5%) HCW-swabs based on RNase P RNA detection (p=0.24). Among symptomatic children, there was no difference in specimen adequacy overall or stratified by age. However, one or both swabs from 19 participants would have been considered inadequate based on RNase P RNA detection alone: 9 self-swabs, 9 HCW-swabs, and 1 both swabs (Figure S2). For asymptomatic children, all HCW-collected and 83 self-collected swabs were considered adequate (p=0.01; Table 3, Figure S3A). RNase P RNA Ct values were similar for children who had adequate self- and HCW-swabs (Figures S2C and S3B). A single individual had detectable SARS-CoV-2 RNA, which was detected in both the self- and HCW-collected swab. This result was confirmed in the CDC EUA rRT-PCR. Notably, 4 individuals with inadequate self-swabs on the first attempt were re-contacted and were trained again on self-swabbing. All self-swabs from the repeat collection had detectable RNase P RNA.

**Table 3.** Characteristics of asymptomatic children categorized based on concordant and discordant results for investigational RNase P RNA detection between self- and HCW-collected AN swabs.

| Category, n (%) | Self-Collection Results |  |  | P-Value |
| --- | --- | --- | --- | --- |
|  | All Participants<br>N=90 | Concordant<br>N=83 | Discordant<br>N=7 |  |
| Age |  |  |  | 0.672 |
| ≤5 | 11 (12.2%) | 10 (12.0%) | 1 (14.3%) |  |
| 6 | 9 (10.0%) | 8 (9.6%) | 1 (14.3%) |  |
| 7 | 13 (14.4%) | 12 (14.5%) | 1 (14.3%) |  |
| 8 | 10 (11.1%) | 10 (12.0%) | 0 (0.00%) |  |
| 9 | 14 (15.6%) | 12 (14.5%) | 2 (28.6%) |  |
| 10 | 13 (14.4%) | 12 (14.5%) | 1 (14.3%) |  |
| 11 | 15 (16.7%) | 15 (18.1%) | 0 (0.00%) |  |
| 12 | 5 (5.6%) | 4 (4.8%) | 1 (14.3%) |  |
| Q4. Did the participant require assistance? |  |  |  | 1.000 |
| No | 79 (87.8%) | 73 (88.0%) | 6 (85.7%) |  |
| Yes | 11 (12.2%) | 10 (12.0%) | 1 (14.3%) |  |
| Q5. Was the swab collection completed correctly? |  |  |  | 0.673 |
| No | 28 (31.1%) | 25 (30.1%) | 3 (42.9%) |  |
| Yes | 62 (68.9%) | 58 (69.9%) | 4 (57.1%) |  |

Finally, 21 samples were tested for Spike receptor binding domain mutations; 18/21 (85.7%) had interpretable results and had evidence of infection with delta variant (K417, L452R, T478K).

### Usability analysis

Children ≤8 years-old were more likely than those > 8 years-old to be scored as having difficulty performing AN swab collection [37/134 (27.6%) vs. 19/147 (12.9%), respectively; p=0.003] and requiring assistance [29/133 (21.8%) vs. 9/147 (6.1%); p<0.001], and they were less likely to be rated as having completed sample collection correctly [82/133 (61.6%) vs. 122/146 (83.6%); p<0.001; Table S2 and S3]. The most common type of assistance provided was instructional regarding how to swab the nostrils. Despite these findings, the proportion of adequate samples, defined as either SARS-CoV-2 RNA detection in the CDC EUA rRT-PCR or RNase P RNA detection, was similar for asymptomatic and symptomatic children ≤8 and >8 years old.

## Discussion

The COVID-19 pandemic has illuminated the need for increased testing capacity across all age groups. Although SARS-CoV-2 testing capacity has dramatically increased, namely due to new testing technologies receiving EUA and deployed for lab and non-lab settings, many testing schemas involve HCWs for sampling. There have been several tests and collection kits deployed for use in the home which utilize self-collection and collection from children by their parents or guardians, but few options exist for pediatric self-collection. This has made it challenging to test and screen pediatric populations due to the need for trained individuals at the school or group setting to collect samples. One prior study investigated the feasibility of AN swab self-collection by school-aged children based on the rate of observed deviations from a standard collection protocol, but results of molecular testing were not reported.^9^ In contrast, the studies presented here characterized the ability of school-aged children to adequately self-collect AN samples based on the results of SARS-CoV-2 molecular testing and evaluated the useability of self-collection from both the child and HCW perspective.

When provided with video and printed instructions, children in all tested age groups were able to adequately self-collect AN samples for the purposes of SARS-CoV-2 testing. There was no significant bias or improvement in performance based on collected demographic variables. These findings support that pediatric self-collection can be used for SARS-CoV-2 testing and could decrease the burden of HCW requirements in existing testing strategies. Additionally, the results support the potential for non-traditional testing schemas for children, including self-collection and sample drop off at schools, prior to events, and even testing at home. These results fill an important void and contribute to the scarce data that exists on pediatric involvement in ongoing testing efforts.

This study was planned and initiated during a nadir in COVID-19 cases in Georgia. Detection of SARS-CoV-2 RNA was the preferred endpoint, but with low rates of infection during the planning phase of the study, additional exploratory outcome measures were considered. As such, RNase P RNA was evaluated as an ancillary measure of specimen adequacy. This required an additional step following nucleic acid extraction to remove genomic DNA, which is also detected in the RNase P assay that is commonly used as a specimen control.^5^ RNase P RNA was detectable in the majority of AN swabs, with no significant difference between self- and HCW-collected samples. One or both swabs from 19 symptomatic and 7 self-swabs from asymptomatic participants were considered inadequate based on RNase P RNA detection. However, no meaningful difference in SARS-CoV-2 RNA detection using the CDC EUA rRT-PCR was detected, which was the primary outcome measure of the study. These data indicate that, although RNase P RNA detection provides a rigorous indicator of specimen adequacy, it may be a more demanding measure than viral RNA detection and overestimate collection failure.

There are a few limitations to the work presented. Data comparing SARS-CoV-2 detection in self-versus HCW-collected samples were limited to symptomatic participants, and there was not sufficient statistical power to detect small differences in SARS-CoV-2 detection by year of age. There was only one asymptomatic child who tested positive for SARS-CoV-2, which did result in positive results for both self- and HCW-collected samples. Further testing should be done to confirm that self-collection is effective for SARS-CoV-2 testing in an asymptomatic pediatric population.

Given the similar performance of self- and HCW-collected AN swabs, data from the current study support the development of SARS-CoV-2 testing plans for school aged children that allow for self-collection. Such strategies have the potential to improve testing capacity by decreasing the need for trained staff at collection sites, and future work could evaluate the use of self-collected samples for multiplex respiratory viral testing to reduce outbreaks in this population.

## Data Availability

All data produced in the present study are available upon reasonable request to the authors

## Acknowledgements

The authors thank the participants and their families for participation in this study. We acknowledge the contributions of Children’s Healthcare of Atlanta Research Coordinators Misty McGhee, Lauren Robinsons, and Rashad Wood on this project. We also acknowledge the helpful comments and feedback from Emory RADx team members during all phases of the study. The Children’s Clinical and Translational Discovery Core is generously supported by Children’s Healthcare of Atlanta and Emory University.

