## Supplemental Material for "Adequacy of Nasal Self-Swabbing for SARS-CoV-2 Testing in Children"

**Adequacy of Self-Collected Anterior Nares Swabs for SARS-CoV-2 Testing by Grade School Children**

### Methods

#### Self-swab evaluation questionnaire

*To participant:* What did you think about the video?

1. Enter participant response ________________

2. Did the participant attempt the swab collection? **(Y/N)**

3. Did you observe the participant experiencing significant difficulties while completing the swab collection? **(Y/N)**

4. Did the participant require assistance? **(Y/N)**
   **à** If yes, what kind of assistance? **(Technical, Instructional, Task)**
   - During which task(s) did the participant require assistance? **(Opening the packaging, grasping the swab, swabbing the nostrils, placing swab in collection tube)**

5. Was the swab collection completed correctly? **(Y/N)**

*To participant:* Can you tell me what it was like to use the swab in your nose?

6. Sum up participant response *(e.g., "scary," "easy," "hard," "gross," "fun," etc.)*___________

*To participant:* Can you tell me how it felt to use the swab in your nose?

7. Enter participant response __________

*To participant:* Was there anything that was hard to do?

8. *Use participant response to answer the following question:* Did the participant indicate difficulties completing the swab collection? **(Y/N/Unsure)**
   **à** If yes During which task(s) did the participant indicate difficulty? **(Opening the packaging, grasping the swab, swabbing the nostrils, placing swab in collection tube)**

*To participant:* Do you think your friends in the same grade as you would be able to use the swab in their own nose?

9. Enter participant response **(Y/N)**

*To participant:* Can you pretend I've never used a swab in my nose and explain to me how to do it?

10. Did participant accurately explain all steps? **(Y/N)**
11. Record any other relevant comments from participant ______________

**Self-collection Instructional Handout: Images and Written Instructions**

**Step 1:** Wash hands with soap and water, or use hand sanitizer.

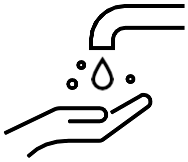

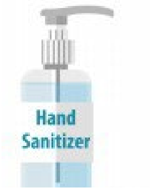


**Step 2:** When instructed by your teacher to do so, open the tube containing the swab. Be sure to only touch the swab handle, not the soft tip of the swab. Do not let the swab touchanything. If the swab falls or touches anything prior to step 3, please let your teacher know and ask for a new swab.

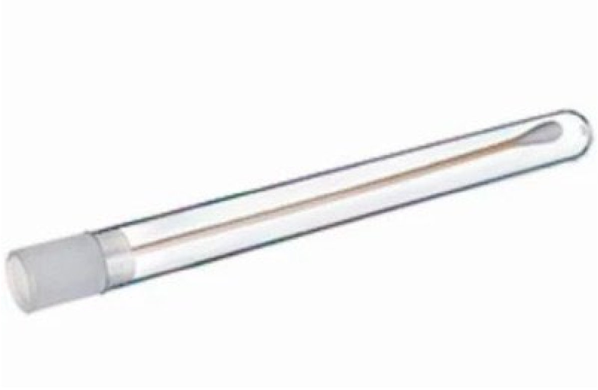

**Step 3:** Gently insert the entire soft tip of the swab into one nostril, and rub the swab slowly in a circular motion around theinside wall of your nostril four times. The swab tip should be touching the inside wall of the nostril through each rotation. Repeat the same process with the same swab on the othernostril.

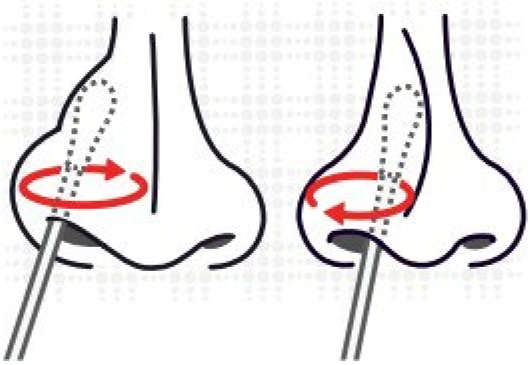

**Step 4:** Insert the swab back into the tube provided, or placethe swab into a container as directed by your teacher.

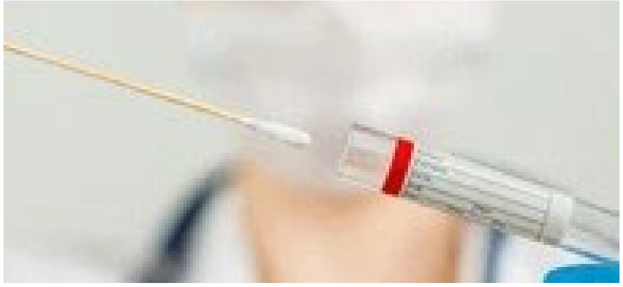

**Self-collection Instructional Handout: Images**

Step 1:

Clean Hands

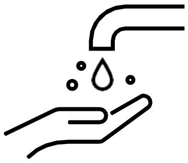

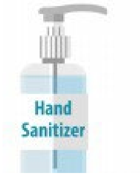


Step 2:

Open Tube

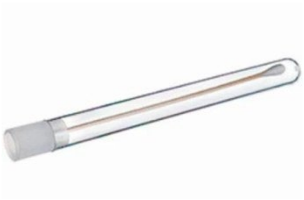

Step 3:

Swab Front of Nose 4 Times on Each Side

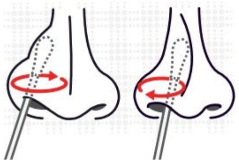

Step 4:

Put Swab Back in Tub or Cup

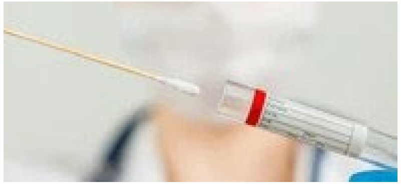

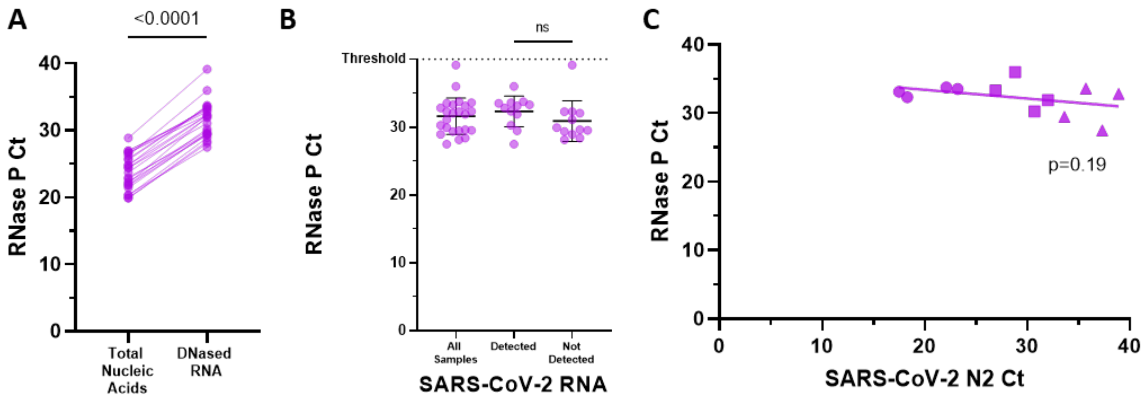


**Figure S1. RNase P RNA is detectable in anterior nares (AN) swabs from asymptomatic children with and without detectable SARS-CoV-2 RNA. A)** DNase treatment of total nucleic acid eluates significantly increased RNase P Ct values (mean difference, 7.8 cycles; standard deviation, 1.2). **B)** RNase P RNA was detectable in all AN swabs, and RNase P Cts did not differ significantly between individuals with and without detectable SARS-CoV-2 RNA. **C)** No trend in RNase P Ct was observed based on SARS-CoV-2 Ct among samples with low (<24, ●), medium (24-32, ■), and high (>32, ▲) Ct values.

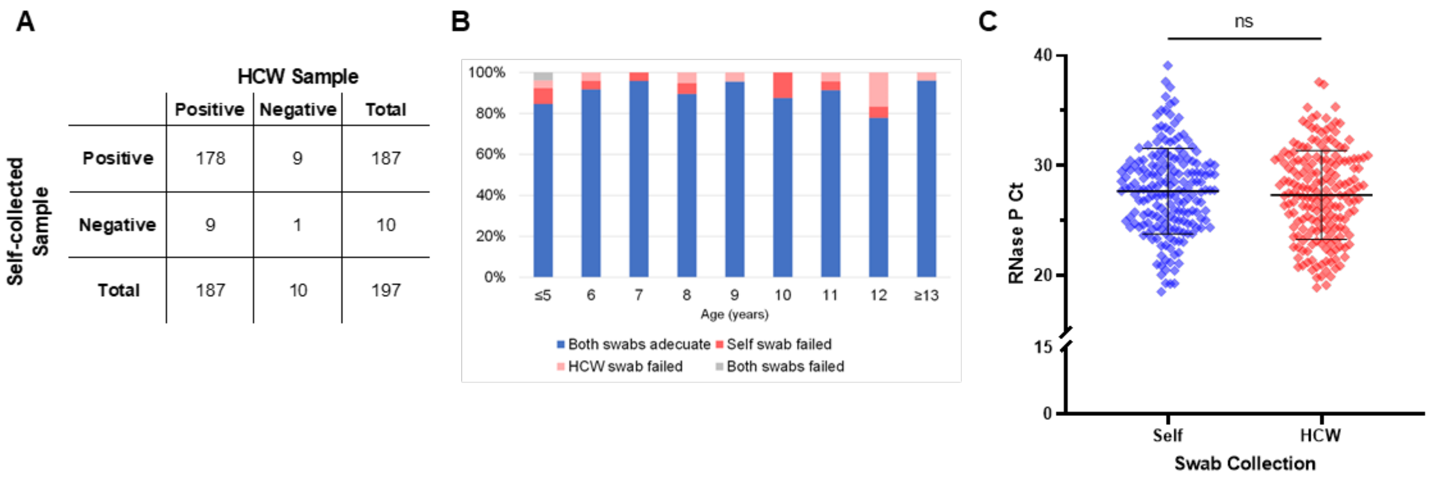


**Figure S2. Collection of adequate specimens did not differ for self- and HCW-collected swabs in symptomatic children when tested for RNase P RNA. A**) 2x2 comparison of qualitative RNase P RNA detection for self- and HCW-collected swabs. **B**) Adequacy of swab pairs by participant age. No significant differences in swab adequacy were observed between the age categories. C) RNase P Ct values for self- and HCW-collected swabs.

A

| Self-collected Sample | HCW Sample: Positive | HCW Sample: Negative | HCW Sample: Total |
|---|---|---|---|
| Positive | 83 | 0 | 83 |
| Negative | 7 | 0 | 7 |
| Total | 90 | 0 | 90 |

B

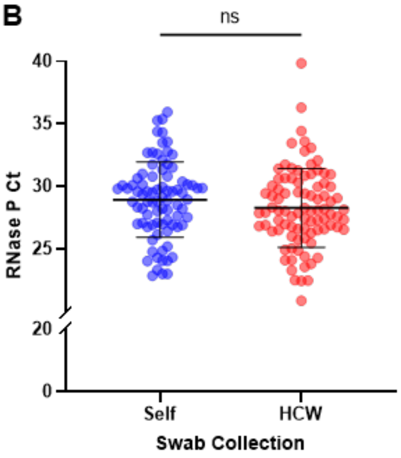


**Figure S3. Comparison of self- versus HCW-collected nasal swab adequacy in asymptomatic children based on RNase P RNA detection. A**) 2x2 comparison of qualitative RNase P RNA detection and **B**) distribution of RNase P Ct values for self- and HCW-collected swabs.

**Table S1.** Analysis of the direction and degree of SARS-CoV-2 N2 Ct differences in the CDC EUA rRT-PCR between self- and HCW-collected swabs.

| Category, n (%) | <3.3 cycles N=59 | Self-collected ≥ 3.3 cycles lower N=12 | HCW-collected ≥ 3.3 cycles lower N=16 | P-Value |
|---|---|---|---|---|
| Gender | | | | 0.940 |
| Female | 28 (48.3%) | 6 (50.0%) | 8 (53.3%) | |
| Male | 30 (51.7%) | 6 (50.0%) | 7 (46.7%) | |
| Race/ethnicity | | | | **0.004** |
| White | 17 (30.4%) | 1 (8.3%) | 8 (53.3%) | |
| Black/African American | 37 (66.1%) | 10 (83.3%) | 4 (26.7%) | |
| Hispanic | 2 (3.6%) | 0 (0.0%) | 1 (6.7%) | |
| Other | 0 (0.0%) | 1 (8.3%) | 2 (13.3%) | |
| Age in years, median (IQR) | 9 (7, 12) | 10 (8, 11) | 9 (7.5, 11) | 0.619 |
| Age Group | | | | 0.288 |
| ≤5 | 7 (11.9%) | 0 (0.0%) | 1 (6.3%) | |
| 6 | 7 (11.9%) | 0 (0.0%) | 2 (12.5%) | |
| 7 | 6 (10.2%) | 3 (25.0%) | 1 (6.3%) | |
| 8 | 8 (13.6%) | 0 (0.0%) | 1 (6.3%) | |
| 9 | 7 (11.9%) | 2 (16.7%) | 4 (25.0%) | |
| 10 | 3 (5.1%) | 4 (33.3%) | 2 (12.5%) | |
| 11 | 6 (10.2%) | 0 (0.0%) | 3 (18.8%) | |
| 12 | 6 (10.2%) | 1 (8.3%) | 1 (6.3%) | |
| ≥13 | 9 (15.3%) | 2 (16.7%) | 1 (6.3%) | |
| Days post-symptom onset, median (IQR) | 3 (2, 4) | 1 (1, 3) | 4 (1, 4) | **0.036** |
| Q4. Did the participant require assistance? | | | | 0.245 |
| No | 52 (89.7%) | 10 (90.9%) | 11 (73.3%) | |
| Yes | 6 (10.3%) | 1 (9.1%) | 4 (26.7%) | |
| Q5. Was the swab collection completed correctly? | | | | 0.268 |
| No | 16 (27.6%) | 7 (46.7%) | 2 (18.2%) | |
| Yes | 42 (72.4%) | 8 (53.3%) | 9 (81.8%) | |

* 2 participants missing gender, 4 missing race/ethnicity, 7 missing days post-symptom onset, 3 missing Q4 and Q5

**Table S2.** Complete useability questionnaire results from symptomatic children, stratified by age.

| Category, n | Age ≤5 | 6 | 7 | 8 | 9 | 10 | 11 | 12 | ≥13 |
|---|---|---|---|---|---|---|---|---|---|
| Q1. HCW to participant: What did you think about the video? | | | | | | | | | |
| Contained “good” | 5 | 8 | 4 | 5 | 7 | 5 | 9 | 7 | 7 |
| Contained “easy” | 3 | 3 | 6 | 2 | 2 | 0 | 3 | 2 | 5 |
| Contained “liked it” | 4 | 2 | 6 | 1 | 4 | 1 | 0 | 0 | 0 |
| Q2. Did the participant attempt the swab collection? | | | | | | | | | |
| No | 1 | 0 | 0 | 0 | 0 | 0 | 0 | 0 | 0 |
| Yes | 23 | 24 | 24 | 19 | 21 | 16 | 23 | 18 | 23 |
| Q3. Did you observe the participant experiencing significant difficulties while completing the swab collection? | | | | | | | | | |
| No | 12 | 20 | 17 | 19 | 18 | 14 | 18 | 14 | 21 |
| Yes | 12 | 4 | 7 | 0 | 3 | 2 | 4 | 4 | 2 |
| Q4. Did the participant require assistance? | | | | | | | | | |
| No | 15 | 19 | 19 | 18 | 19 | 14 | 18 | 18 | 23 |
| Yes | 9 | 5 | 4 | 1 | 2 | 2 | 4 | 0 | 0 |
| Q4a. If yes, what kind of assistance? | | | | | | | | | |
| Technical | 2 | 3 | 3 | 0 | 2 | 2 | 2 | 0 | 0 |
| Instructional | 7 | 2 | 3 | 1 | 0 | 0 | 3 | 0 | 0 |
| Task | 2 | 0 | 2 | 1 | 0 | 0 | 0 | 0 | 0 |
| Q4b. During which task(s) did the participant require assistance? | | | | | | | | | |
| Opening the packaging | 2 | 1 | 2 | 0 | 2 | 2 | 2 | 0 | 0 |
| Grasping the swab | 1 | 0 | 2 | 0 | 0 | 0 | 0 | 0 | 0 |
| Swabbing the nostrils | 7 | 4 | 2 | 1 | 0 | 1 | 3 | 0 | 0 |
| Other | 1 | 0 | 1 | 0 | 0 | 0 | 0 | 0 | 0 |
| Q4 Other. During which task(s) did the participant require assistance? | | | | | | | | | |
| Afraid to swab herself | 0 | 0 | 1 | 0 | 0 | 0 | 0 | 0 | 0 |
| Afraid to test himself. Received NP day before. | 1 | 0 | 0 | 0 | 0 | 0 | 0 | 0 | 0 |
| Q5. Was the swab collection completed correctly? | | | | | | | | | |
| No | 13 | 9 | 4 | 4 | 5 | 4 | 2 | 3 | 3 |
| Yes | 11 | 14 | 20 | 15 | 16 | 12 | 20 | 15 | 19 |
| Q6. HCW to participant: Can you tell me what it was like to use the swab in your nose? | | | | | | | | | |
| Contained “easy” | 5 | 12 | 10 | 9 | 8 | 3 | 11 | 11 | 13 |
| Contained “tickled” | 2 | 1 | 4 | 1 | 3 | 0 | 3 | 0 | 0 |
| Contained “fun” | 1 | 1 | 1 | 2 | 2 | 2 | 0 | 0 | 2 |
| Contained “okay’ | 3 | 2 | 1 | 0 | 1 | 2 | 2 | 0 | 0 |
| Q7. HCW to participant: Can you tell me how it felt to use the swab in your nose? | | | | | | | | | |
| Contained “tickled” | 5 | 10 | 14 | 8 | 7 | 9 | 8 | 6 | 7 |
| Contained “did not hurt” | 2 | 1 | 0 | 0 | 1 | 0 | 1 | 2 | 3 |
| Contained “soft” | 3 | 1 | 0 | 2 | 2 | 0 | 0 | 1 | 0 |
| Q8. HCW to participant: Was there anything that was hard to do? Did the participant indicate difficulties completing the swab collection? | | | | | | | | | |
| No | 18 | 21 | 21 | 18 | 18 | 16 | 21 | 18 | 22 |

| Category, n | ≤5 | 6 | 7 | 8 | Age<br>9 | 10 | 11 | 12 | ≥13 |
|---|---|---|---|---|---|---|---|---|---|
| Yes | 4 | 2 | 1 | 1 | 3 | 0 | 1 | 0 | 1 |
| Unsure | 1 | 1 | 1 | 0 | 0 | 0 | 0 | 0 | 0 |
| Q8a. If yes, during which task(s) did the participant indicate difficulty? | | | | | | | | | |
| Opening the packaging | 1 | 0 | 0 | 0 | 0 | 0 | 1 | 0 | 1 |
| Grasping the swab | 1 | 0 | 0 | 0 | 0 | 0 | 0 | 0 | 0 |
| Swabbing the nostrils | 3 | 2 | 0 | 1 | 2 | 0 | 0 | 0 | 0 |
| Other | 0 | 0 | 1 | 0 | 0 | 0 | 0 | 0 | 0 |
| Q8a Other. Describe other | | | | | | | | | |
| Complete swabbing | 0 | 0 | 1 | 0 | 0 | 0 | 0 | 0 | 0 |
| Q9. HCW to participant: Do you think your friends in the same grade as you would be able to use the swab in their own nose? | | | | | | | | | |
| No | 2 | 3 | 1 | 1 | 2 | 0 | 1 | 1 | 0 |
| Yes | 19 | 20 | 21 | 18 | 19 | 16 | 21 | 17 | 23 |
| Q10. HCW to participant: Can you pretend I've never used a swab in my nose and explain to me how to do it? | | | | | | | | | |
| No | 14 | 10 | 4 | 2 | 5 | 0 | 2 | 2 | 3 |
| Yes | 8 | 14 | 19 | 17 | 16 | 16 | 20 | 16 | 20 |

* 6 participants missing Q1, 5 missing Q2, 6 missing Q3, 7 missing Q4, 5 missing Q4a, 5 missing Q4b, 6 missing Q6, 6 missing Q7, 8 missing Q8, 5 missing Q8a, 12 missing Q9, 9 missing Q10

**Table S3.** Complete useability questionnaire results from asymptomatic children, stratified by age.

| Category, n | Age | | | | | | | |
|---|---|---|---|---|---|---|---|---|
| | ≤5 | 6 | 7 | 8 | 9 | 10 | 11 | 12 |
| Q1. HCW to participant: What did you think about the video? | | | | | | | | |
| Contained "good" | 3 | 8 | 6 | 5 | 3 | 3 | 3 | 1 |
| Contained "helpful" | 0 | 0 | 3 | 2 | 4 | 4 | 6 | 0 |
| Contained "okay" | 2 | 1 | 0 | 1 | 2 | 3 | 2 | 2 |
| Q2. Did the participant attempt the swab collection? | | | | | | | | |
| No | 2 | 0 | 0 | 0 | 0 | 0 | 0 | 0 |
| Yes | 9 | 9 | 13 | 10 | 14 | 13 | 15 | 5 |
| Q3. Did you observe the participant experiencing significant difficulties while completing the swab collection? | | | | | | | | |
| No | 8 | 8 | 5 | 8 | 10 | 13 | 15 | 5 |
| Yes | 3 | 1 | 8 | 2 | 4 | 0 | 0 | 0 |
| Q4. Did the participant require assistance? | | | | | | | | |
| No | 6 | 8 | 9 | 10 | 14 | 13 | 14 | 5 |
| Yes | 5 | 1 | 4 | 0 | 0 | 0 | 1 | 0 |
| Q4a. If yes, what kind of assistance? | | | | | | | | |
| Technical | 2 | 0 | 2 | 0 | 0 | 0 | 0 | 0 |
| Instructional | 5 | 1 | 2 | 0 | 0 | 0 | 1 | 0 |
| Task | 1 | 1 | 2 | 0 | 0 | 0 | 0 | 0 |
| Q4b. During which task(s) did the participant require assistance? | | | | | | | | |
| Opening the packaging | 1 | 1 | 0 | 0 | 0 | 0 | 0 | 0 |
| Grasping the swab | 1 | 1 | 1 | 0 | 0 | 0 | 0 | 0 |
| Swabbing the nostrils | 5 | 1 | 4 | 0 | 0 | 0 | 1 | 0 |
| Other | 1 | 0 | 0 | 0 | 0 | 0 | 0 | 0 |
| Q4 Other. During which task(s) did the participant require assistance? | | | | | | | | |
| Touching the swab | 1 | 0 | 0 | 0 | 0 | 0 | 0 | 0 |
| Q5. Was the swab collection completed correctly? | | | | | | | | |
| No | 8 | 3 | 8 | 2 | 3 | 2 | 2 | 0 |
| Yes | 3 | 6 | 5 | 8 | 11 | 11 | 13 | 5 |
| Q6. HCW to participant: Can you tell me what it was like to use the swab in your nose? | | | | | | | | |
| Contained "easy" | 2 | 4 | 11 | 4 | 8 | 10 | 12 | 2 |
| Contained "tickled" | 2 | 2 | 1 | 1 | 2 | 2 | 0 | 0 |
| Contained "okay" | 1 | 1 | 0 | 0 | 0 | 1 | 1 | 2 |
| Q7. HCW to participant: Can you tell me how it felt to use the swab in your nose? | | | | | | | | |
| Contained "tickled" | 6 | 6 | 9 | 4 | 9 | 5 | 7 | 3 |
| Contained "weird" | 0 | 0 | 2 | 1 | 1 | 4 | 2 | 1 |
| Contained "okay" | 1 | 3 | 0 | 1 | 0 | 0 | 1 | 0 |
| Q8. HCW to participant: Was there anything that was hard to do? Did the participant indicate difficulties completing the swab collection? | | | | | | | | |
| No | 10 | 8 | 11 | 9 | 14 | 13 | 14 | 5 |
| Yes | 1 | 1 | 2 | 1 | 0 | 0 | 0 | 0 |
| Unsure | 0 | 0 | 0 | 0 | 0 | 0 | 1 | 0 |

| Category, n | Age ≤5 | 6 | 7 | 8 | 9 | 10 | 11 | 12 |
|---|---|---|---|---|---|---|---|---|
| Q8a. If yes, during which task(s) did the participant indicate difficulty? | | | | | | | | |
| Opening the packaging | 0 | 0 | 0 | 0 | 0 | 0 | 0 | 0 |
| Grasping the swab | 0 | 0 | 0 | 0 | 0 | 0 | 0 | 0 |
| Swabbing the nostrils | 1 | 1 | 0 | 1 | 0 | 0 | 0 | 0 |
| Other | 0 | 0 | 2 | 0 | 0 | 0 | 0 | 0 |
| Q8a Other. Describe other | | | | | | | | |
| Getting started | 0 | 0 | 1 | 0 | 0 | 0 | 0 | 0 |
| Swabbing himself | 0 | 0 | 1 | 0 | 0 | 0 | 0 | 0 |
| Q9. HCW to participant: Do you think your friends in the same grade as you would be able to use the swab in their own nose? | | | | | | | | |
| No | 2 | 0 | 2 | 1 | 0 | 0 | 1 | 0 |
| Yes | 9 | 9 | 11 | 9 | 14 | 13 | 14 | 5 |
| Q10. HCW to participant: Can you pretend I've never used a swab in my nose and explain to me how to do it? | | | | | | | | |
| No | 7 | 6 | 3 | 1 | 1 | 0 | 2 | 0 |
| Yes | 4 | 3 | 10 | 9 | 13 | 13 | 13 | 5 |
